# Granger causality connectivity analysis of persistent atrial fibrillation dynamics reveals mechanistic insights into the posterior wall

**DOI:** 10.1101/2024.11.11.24317135

**Authors:** Joseph Barker, Arunashis Sau, Nikesh Bajaj, Alex Jenkins, Alex Sharp, Xili Shi, Xinyang Li, Nabeela Karim, Balvinder Handa, Richard Chambers, Timothy Betts, Nicholas S Peters, Tom Wong, Fu Siong Ng

**Affiliations:** National Heart and Lung Institute, Imperial College London, United Kingdom; Department of Cardiology, Imperial College Healthcare NHS Trust, London, United Kingdom; Institute of Biomechanical Engineering, Department of Engineering Science, University of Oxford, Oxford, United Kingdom; Department of Cardiology, Royal Brompton & Harefield Hospitals, Guy’s and St. Thomas’ NHS Foundation Trust, London, United Kingdom; Acutus Medical, Carlsbad, California, USA; ChamberTech LTD, London Institute for Healthcare Engineering, London, United Kingdom; Division of Cardiovascular Medicine, University of Oxford, Oxford, United Kingdom; Department of Cardiology, Chelsea and Westminster Hospital NHS Foundation Trust, London, United Kingdom

## Abstract

**Background:** Pulmonary vein isolation (PVI) is the mainstay of ablation for atrial fibrillation (AF). Adjunctive posterior wall isolation (PWI) has not demonstrated convincing additional benefit. To provide mechanistic underpinnings as to why empirical PWI does not improve outcomes, we undertook Granger Causality (GC) analysis of patient-specific AF dynamics before and after ablation.

**Methods:** A prospective cohort study was undertaken at Royal Brompton Hospital. Consecutive patients undergoing PVI with left atrial electro-anatomical noncontact mapping (AcQmap; Acutus Medical) were included. GC analysis was undertaken before and after PVI but before adjunctive ablation.

**Results:** In 21 consecutive patients, Causality Pairing Index, a Granger Causality-based measure of AF organisation, was unchanged post PVI; overall 0.087±0.012 vs. 0.086±0.015, p = 0.64, or by region (posterior wall; 0.084±0.020 vs 0.079±0.017, p = 0.20, rest of LA 0.087±0.013 vs 0.086±0.016, p = 0.80). Directional dispersion, quantifying conduction heterogeneity, was lower in the PW compared to the rest of the LA (0.093±0.036 vs 0.11±0.043, p = 0.017) and increased following PVI (0.093±0.036 vs 0.12±0.043, p = 0.045), while there was no change in the rest of the LA (0.11±0.034 vs 0.11±0.030, p =0.52). Net outflow for left atrial posterior wall decreased following PVI (pre −0.0086±0.047 vs −0.033±0.054, p=0.011), suggesting that in the majority of cases the posterior wall becomes a net sink after PVI

**Conclusion:** We describe the first application of GC to global, simultaneous AF mapping data. GC analysis suggests, on average, the posterior wall is a net sink following PVI, and therefore PWI will not be beneficial in the majority of patients, providing mechanistic insight into null randomised control trials for PWI. GC is positioned as a valuable clinical decision tool to select the minority of patients that may benefit from PWI to guide personalised PsAF ablation strategies.

**Clinical Perspective:** *What is Known:*

- Pulmonary vein isolation (PVI) is a standard treatment for atrial fibrillation (AF) ablation, but adjunctive posterior wall isolation (PWI) has not demonstrated consistent additional clinical benefit.

*What the Study Adds:*

- Granger Causality analysis of AF dynamics indicates that, following PVI, the left atrial posterior wall becomes a net sink in most cases.
- This finding provides mechanistic insight into why PWI does not improve outcomes for most patients, explaining the null results in randomised trials.
- The study supports using Granger Causality as a decision tool to personalise ablation strategies, identifying the minority of patients who may benefit from PWI.

**Graphical Abstract:** 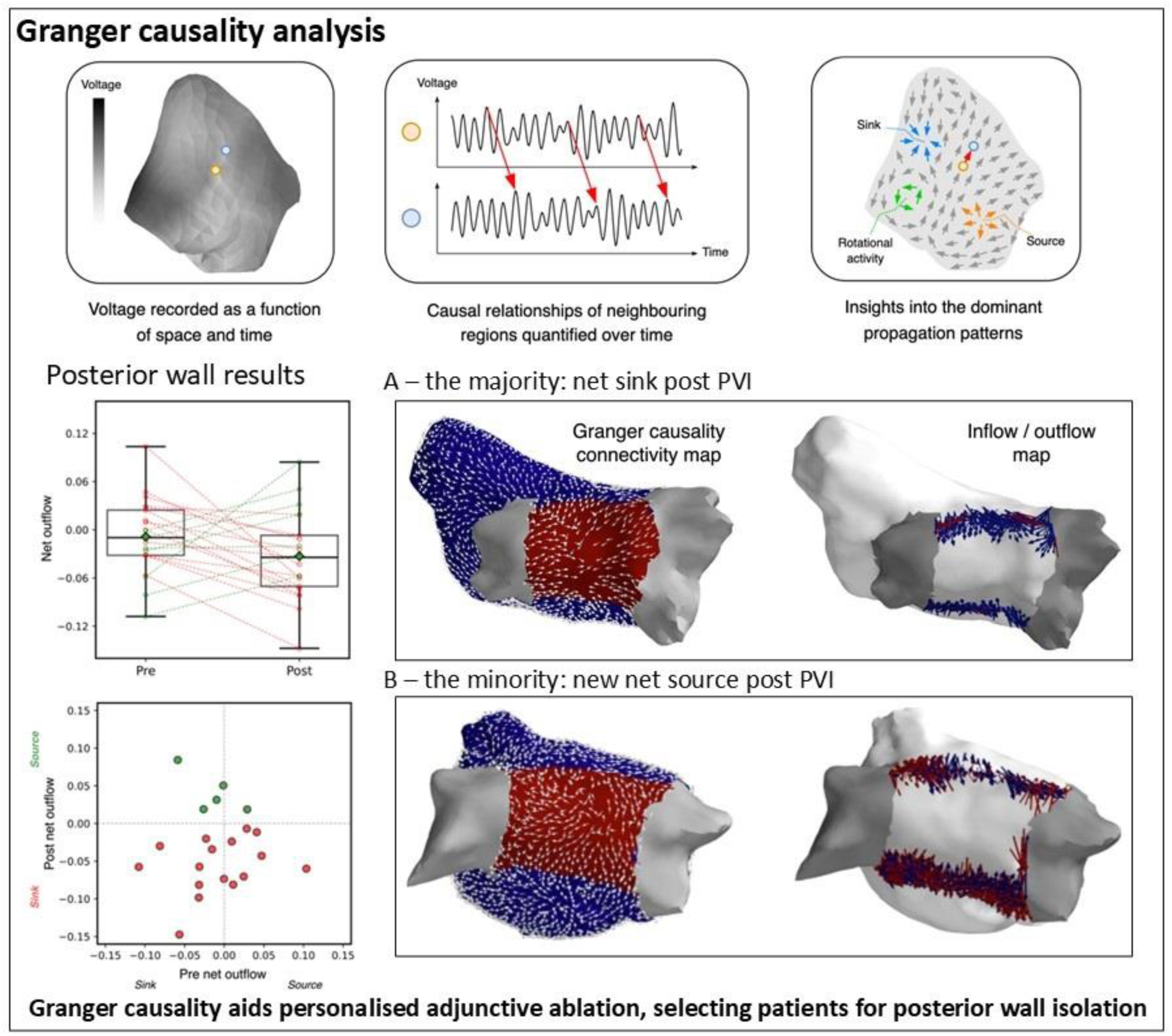

## Introduction

Pulmonary vein isolation (PVI) has been the mainstay of ablation for atrial fibrillation (AF) since its seminal description in 1998 (1). It works by isolating the atria from pulmonary vein ectopy and may result in atrial substrate modification (2–4). PVI for paroxysmal AF (pAF) has good success rates (>70%) whilst PVI for persistent AF (PsAF) has more modest success rates with a ceiling of around 50% (4–6).

Many adjunctive lesion sets have been studied to improve success of PsAF ablation, however none have demonstrated convincing additional benefit beyond PVI alone (6). Posterior wall isolation (PWI), designed to isolate posterior wall sources of ectopy, initially showed promise in the literature. However, subsequent studies failed to demonstrate clear benefit, with the recent results of the rigorously conducted randomised-control CAPLA study, implementing both lesion specific quality targets and intense/complete follow-up, showing no benefit of PWI+PVI vs PVI alone (7–11). Despite this, PWI remains in the AF catheter ablation consensus statements globally (12, 13).

We previously adapted Granger Causality (GC) analysis, an econometric tool, for use in AF mapping (14). This method analyses the “causal” relationships between AF signals to build a picture of AF connectivity, providing insight into patient-specific AF dynamics. In this study, we performed GC analysis on AF mapping data before and after PVI, to better understand the role of the left atrial posterior wall in AF pathophysiology and to provide some mechanistic underpinnings for the CAPLA study and more broadly the negative results of PWI randomised control trials (6).

## Methods

### Ethical approval

This study was approved by the national ethics committee (National Health Service Health Research Authority). Written individual informed patient consent was obtained.

### Clinical Data Collection

A prospective cohort study was undertaken at Royal Brompton Hospital as previously described (15). Briefly, consecutive patients with PsAF undergoing AF ablation had left atrial electro-anatomical maps collected using the AcQMap noncontact mapping catheter. AcQmap uses ultrasound to reconstruct 3D endocardial anatomy, then creates dipole density maps from unipolar electrograms, allowing for real-time, precise mapping of global cardiac electrical activity (16). The reconstructed unipolar electrograms were used in this analysis. AF data were analysed before and after PVI but before any adjunctive ablations (17). Statistical analysis was undertaken on a complete case basis, with this manuscript prepared according to STROBE guidelines (18).

### Granger Causality Analysis

GC was initially described as an econometric tool to quantify causal dependence between time-series (19). We previously described the application of GC to PsAF (14). GC analysis identifies the causal relationships between pairs of electrograms recorded from different locations within a cardiac chamber. The degree and pattern of causal relationships of electrogram signals within the atrium provides indirect information dominant propagation patterns in AF and the AF electrophenotype, **Figure 1**. For each signal, GC analysis is used as a linear model to predict the future sample values from given past values from nearest nodes within a neighbourhood radius of 10mm and including itself. The linear model is optimised with lasso regression to have sparse weights (a few non-zero weights). Since the model is optimised using all the nearest neighbours, the established connections are considered conditional causal connections. Given the non-stationary nature of signals, applying GC over longer length of signal recording will only reveals stationary connections.

**Figure 1.**
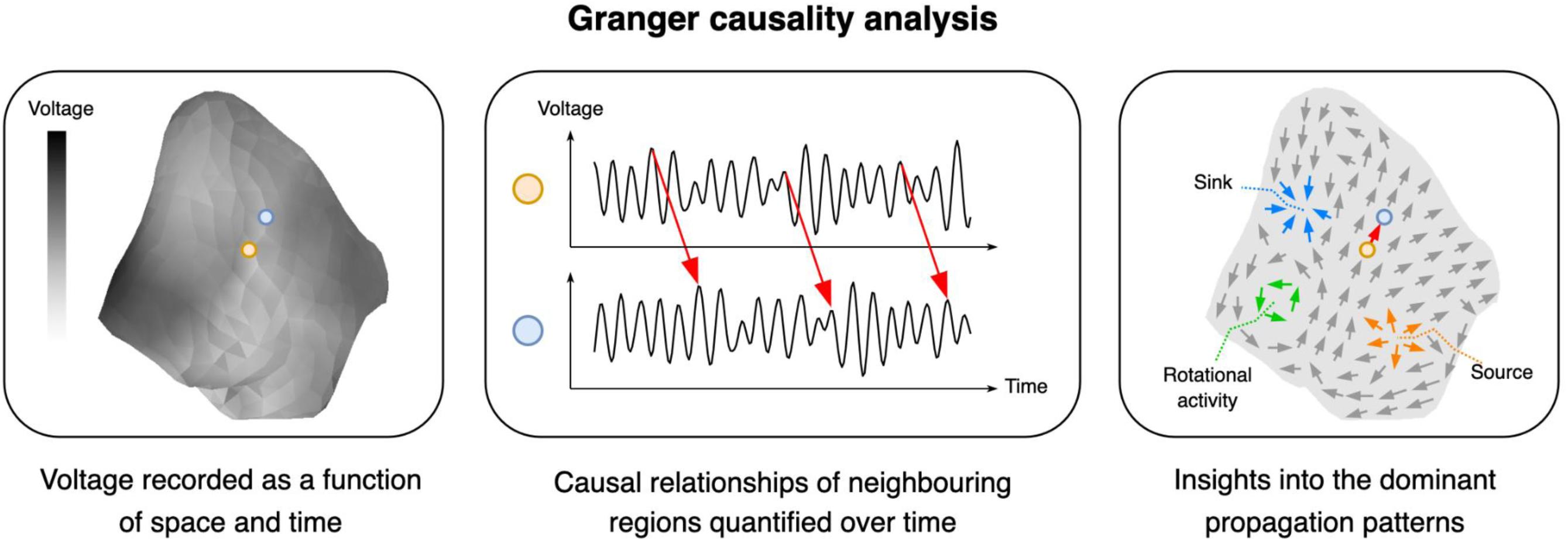
An overview of Granger Causality analysis applied to simultaneous electro-anatomical non-contact mapping

### Parameters to quantify connectivity

We applied three parameters computed using causal connections obtained from conditional GC (20) to describe the fibrillation dynamics in PsAF.

#### (i) Causality Pairing Index

Causality Pairing Index (CPI) is a measure of the degree of connectivity/organisation of the AF. It is computed as a ratio of total number of connections found to total possible connections. It ranges from 0 to 1, where 0 means no connections found and 1 means, every node is connected to every other node. An extreme case of a macro-reentrant tachycardia would be expected to have a CPI close to 1 as the activation at an electrode site can be predicted by an adjacent electrode, given the organised nature of this rhythm. More “organised” AF would be expected to have greater CPI than “disorganised” AF. Further details in **Supplementary Methods** and Supplementary Figure 1.

#### (ii) Directional Dispersion

Directional Dispersion (DD) characterises ‘smoothness’ of flow (21, 22), analogous to conduction vector dispersion, with highly heterogeneous conduction having a high DD. For each signal DD computes the direction of flow and its similarity with direction of neighbouring nodes. DD ranges from 0 and 1, where 0 means the direction of given node is the same as neighbouring nodes and 1 means it is the 180-degree opposite direction. The calculation of DD is described in detail in the **Supplementary Methods** and **Supplementary Figure 2**.

#### (iii Net Outflow

Net outflow for a region is computed as total connections going-out from region subtracted from total connections coming-in (23). This is normalised by total possible connections between region and outside. Net outflow can discern the direction of connections between regions. A positive value of Net outflow for a region indicates region as a source of information flow (which may be a region of an AF driver), while negative value reflects its characteristics to be sink. It ranges from −1 to 1, where −1 indicates all the possible connections from outside of regions are coming-in. Further details described in **Supplementary Methods.**

## Results

A total of 21 participants, with an average age of 64 years and 33% female, were included in the study. Patient charactisterics are presented in **Table 1**. Patients were in persistent AF for an average 9.4 months on 2 antiarrthymic drugs before ablation, with a mean CHAD2S2VASc of 2. The average LA dimension was 43.1mm, with moderate mitral regurgitation in 24% of participants and an average ejection fraction of 49%. Ultimately, 65% of participants were free from AF at 11 months of follow-up.

**Table 1:** Participant characteristics. Mean (SD). Percutaneous coronary intervention (PCI). Left atrium (LA). Left ventricular ejection fraction (LVEF).

| Variable | N = 21 |
| --- | --- |
| Age | 63.8 (11.1) |
| Female % | 7 (33%) |
| Persistent atrial fibrillation % | 21 (100%) |
| Months since diagnosis | 9.4 (5.2) |
| Heart rate | 81.4 (13.8) |
| Systolic blood pressure | 127.0 (17.9) |
| Diastolic blood pressure | 77.0 (11.9) |
| Body mass index | 27.6 (3.7) |
| Hypertension | 9 (43%) |
| Type 2 diabetes mellitus | 4 (19%) |
| Heart failure | 8 (40%) |
| Stroke | 1 (5%) |
| Chronic kidney disease | 0 (0%) |
| Ex-smoker | 9 (43%) |
| Antiarrhythmic drugs (count) | 2 (1) |
| CHAD2S2VASc | 2 (1) |
| LA Diameter mm | 43.1 (5.0) |
| Moderate mitral regurgitation | 5 (24%) |
| LVEF % | 49.2 (13.4) |
| Follow up in months | 11 (5) |
| Freedom from AF at follow up | 65% |

Non-contact mapping was performed in patients undergoing ablation for PsAF. Mapping was performed pre and post PVI. The first five seconds of AF data was used to compute connectivity measures pre- and post-PVI. The LA was divided into two regions, the posterior wall (PW) and the rest of the left atrium, **Figure 2**.

**Figure 2.**
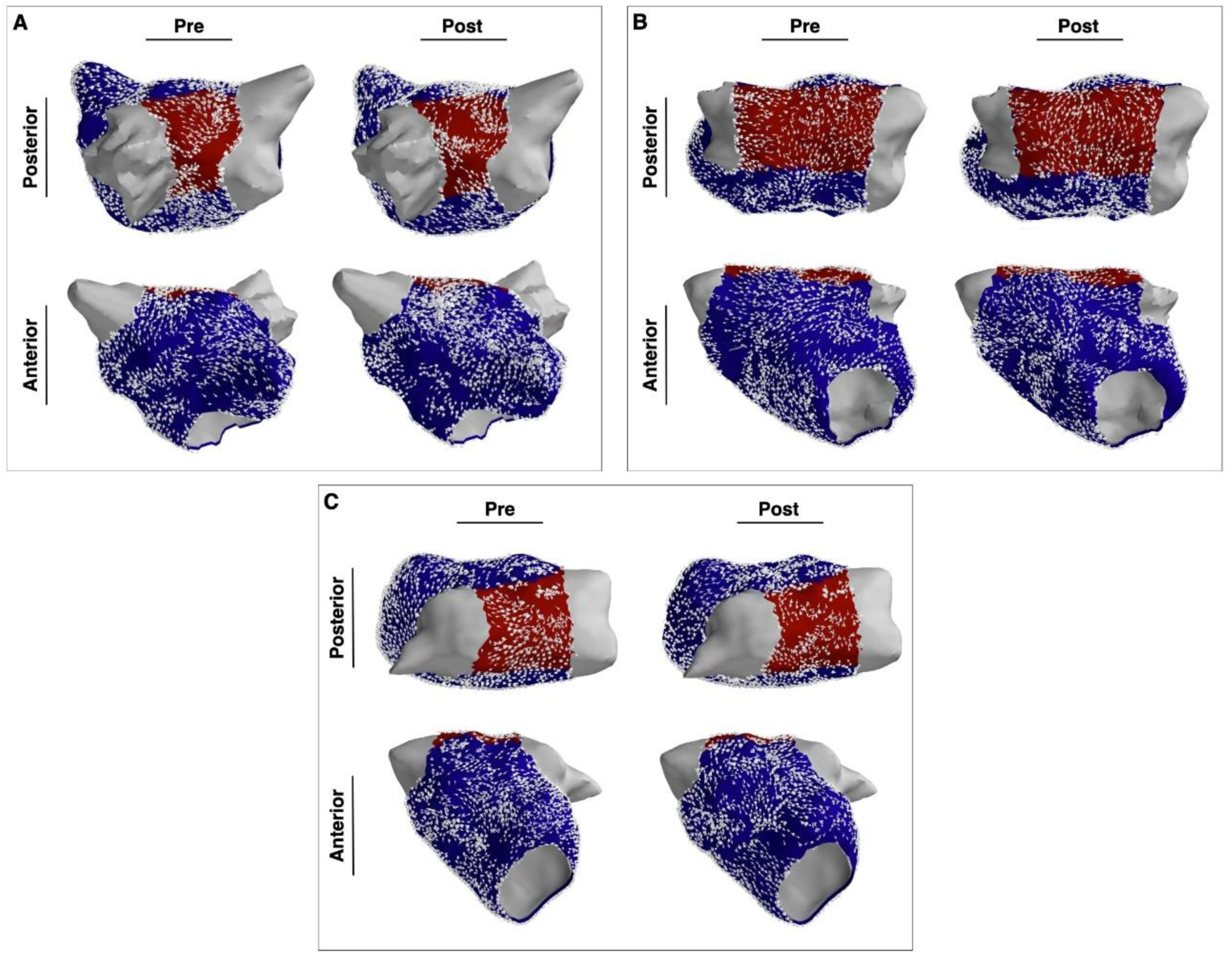
Three representative examples showing left atrial Granger causality (GC) connectivity maps. There is no significant difference in connectivity by region or pre/post pulmonary vein isolation. Top panels show posterior view, bottom panels show an anterior view. The pulmonary veins are depicted in grey, the posterior wall is depicted in red, and the rest of the left atrium is blue

### Causality pairing index

CPI is a measure of organisation, with high CPI indicating more organised fibrillation. **Figure 3** shows examples of GC connectivity maps pre and post PVI. CPI did not significantly change overall post PVI (0.087±0.012 vs. 0.086±0.015, p = 0.64) or by region (PW 0.084±0.020 vs 0.079±0.017, p = 0.20, Rest 0.087±0.013 vs 0.086±0.016, p = 0.80, **Figure 3**).

**Figure 3.**
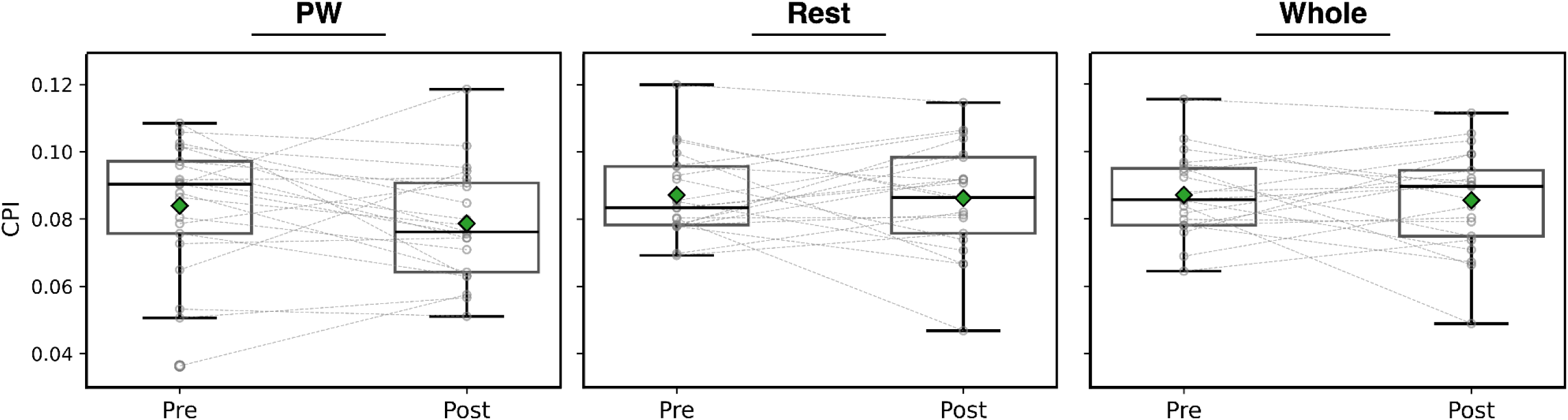
Causality pairing index (CPI) was computed pre and post pulmonary vein isolation (PVI) for the left atrium (LA) as a whole and by region. There were no significant differences in CPI by region or pre/post PVI

### Directional dispersion

Directional dispersion quantifies conduction heterogeneity in each area of interest, higher directional dispersion signifies greater conduction heterogeneity. Prior to PVI, mean directional dispersion mean DD was lower in the PW compared to the rest of the LA (Figure 3, 0.093±0.036 vs 0.11±0.043, p = 0.017). PW directional dispersion increased following PVI (**Figure 4**, 0.093±0.036 vs 0.12±0.043, p = 0.045), while there was no change in the rest of the LA (0.11±0.034 vs 0.11±0.030, p =0.52). **Figure 5** shows examples of low to high and high to low directional dispersion in the PW.

**Figure 4.**
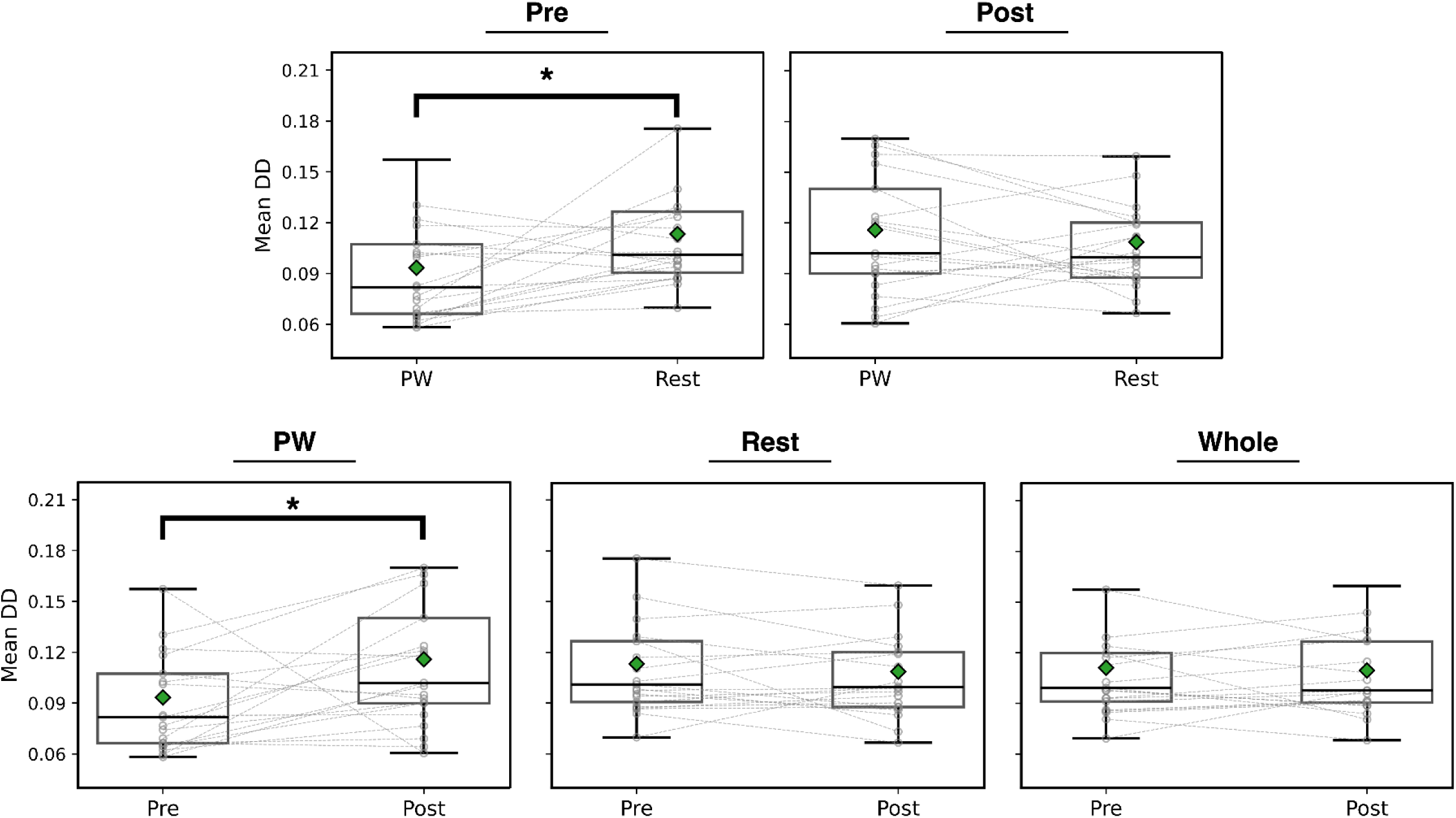
Mean directional dispersion (DD) is a measure of conduction heterogeneity. Mean DD was significantly lower in the posterior wall (PW) compared to the rest of the left atrium (LA) pre pulmonary vein isolation (PVI), but not post ablation. Overall, mean DD in the PW significantly increased following PVI, however there were a minority of cases where the opposite trend is observed.

**Figure 5.**
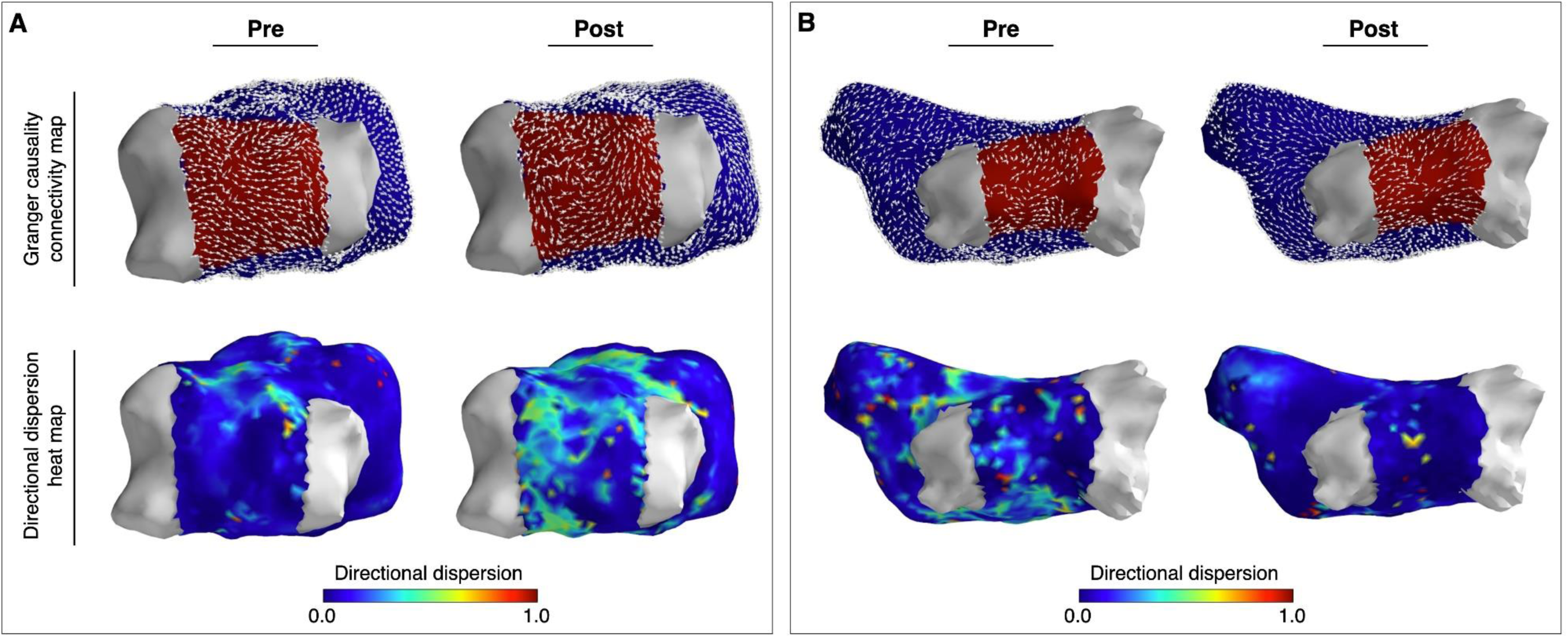
Representative examples showing Granger causality (GC) connectivity maps and directional dispersion heat maps. (A) Directional dispersion (DD) in the posterior wall (PW) increases following pulmonary vein isolation (PVI). (B) DD in the PW decreases following PVI. The pulmonary veins are depicted in grey, the PW is depicted in red, and the rest of the left atrium is blue

### Net Outflow for left atrial posterior wall

In order to understand the significance of the posterior wall as a potential region that may harbour AF drivers following PVI, we computed the net outflow, which describes the overall direction of connections between regions. We found the net outflow of the posterior wall was, on average, zero (i.e. equal connections going into and out of the PW) pre PVI (−0.0086±0.047, null hypothesis - net outflow ≈ 0, one-sample t-test, p = 0.41). However, following PVI, the Net Outflow was, on average, negative (post PVI −0.033±0.054, p = p=0.011), suggesting that in the majority of cases the posterior wall becomes a net sink after PVI. **Figure 6** shows the posterior wall in 16 samples was a net sink post PVI, 5 were a net source. **Figure 7** shows representative examples where the posterior wall is a net sink and net source.

**Figure 6.**
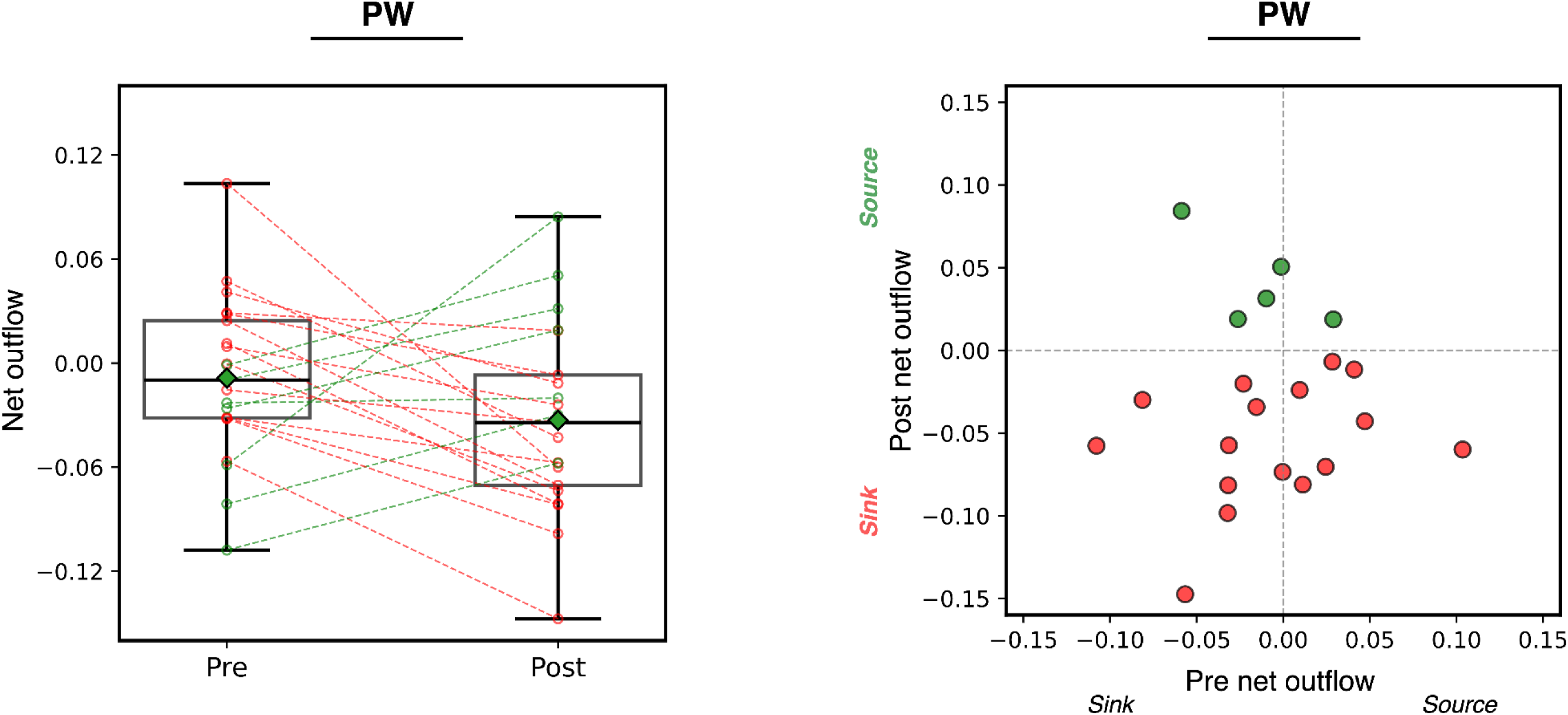
Granger causality (GC) connectivity maps were used to compute net outflow from the posterior wall (PW). Positive net outflow indicates the PW was a net source, while negative indicates a net sink. Overall, there was no significant change in outflow following pulmonary vein isolation (PVI). After PVI, in the majority the PW was a net sink, but was a source in a minority of cases.

**Figure 7.**
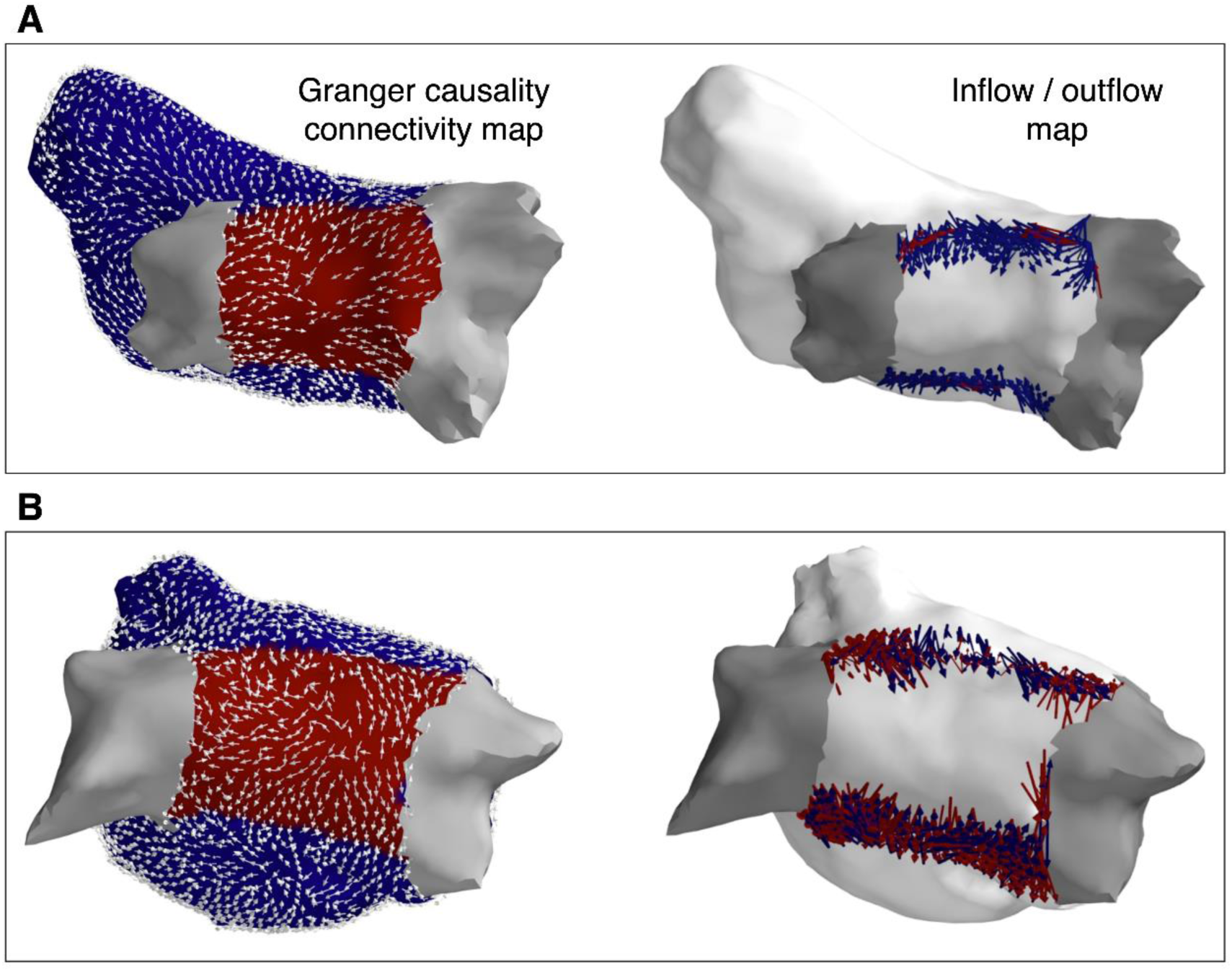
Representative examples showing Granger causality (GC) connectivity maps and posterior wall outflow post PVI. In the inflow/outflow maps blue arrows indicate inflow into the PW and red indicate outflow. (A) example of a net sink, (B) example of a new source. The pulmonary veins are depicted in grey, the PW is depicted in red, and the rest of the left atrium is blue

## Discussion

In this study, we demonstrate the feasibility of Granger Causality to quantify global fibrillation organisation, characterise dominant propagation patterns and identify driver regions (sources) and sinks in simultaneously collected non-contact charge density PsAF maps. We found that overall, the degree of organisation does not change after PVI but that PWI is likely ineffective as an empiric adjunctive procedure because the PW becomes a net sink for atrial activity following PVI, on average.

### Granger Causality applied to non-contact charge density maps

We have previously described and validated fibrillation analysis using GC-based methods in conjunction with optical mapping and sequentially acquired clinical data (10). A major limitation of sequential data collection is the inherently chaotic and dynamic nature of atrial fibrillation (AF), which precludes the “stitching” of regional data collected in sequence and non-simultaneously. Here, we present a global, simultaneous approach, which provides a more accurate representation of AF dynamics.

Our results demonstrate that global AF organisation remains largely unchanged following PVI, contrasting with prior studies that have reported a prolongation of the AF cycle length (AFCL) and a reduction in complex fractionated electrograms, suggesting increased AF organisation following PVI (2, 3, 24) (25). However, we observed notable alterations in the fibrillation dynamics of the posterior wall, specifically an increase in conduction heterogeneity after PVI. This increase in heterogeneity in the PW may be due to PVI reducing the critical mass of the posterior wall (26), which could disrupt potential driver regions responsible for laminar flow with low conduction heterogeneity. These findings challenge the conventional understanding of post-PVI AF behaviour.

### Posterior wall dynamics

Our findings provide insight into the null results observed in the CAPLA trials with PWI for PsAF ablation (8, 10). We identified that only a minority of patients exhibit net positive outflow from the posterior wall (i.e., the posterior wall acts as a source) following pulmonary vein isolation (PVI). On average, the posterior wall functioned as a net sink, suggesting that electrical isolation of this region may offer limited benefit for most patients. In line with the critical mass hypothesis (26), PVI resulting in a reduction in the effective posterior wall area, compartmentalising the region and diminishing net outflow, could explain our findings.

However, it should be noted that in 24% of patients, the posterior became a net source following PVI, highlighting the heterogeneous response to PVI. Our results suggest that these smaller group of patients may benefit from PWI and that adjunctive strategies should be personalised and more targeted to patients most likely to benefit from the additional lesions, rather than be performed in all-comers in a one-size-fits-all manner.

### Limitations

This study has several limitations that warrant consideration. First, the small sample size limits the generalisability of the findings across diverse patient populations.

Additionally, the single-centre nature of the study may introduce centre-specific biases in patient selection, procedural techniques, and outcomes, which could reduce the external validity of our results. The use of non-contact electrograms, though advantageous for global mapping, introduces potential limitations in data accuracy due to signal noise, interpolation and resolution which could affect the precision of the Granger Causality analysis. Granger Causality itself, based on linear and stationary assumptions, may not fully account for the complex, non-linear nature of atrial fibrillation, and future studies might benefit from incorporating non-linear analytical methods. Moreover, the study was limited to pulmonary vein isolation alone without the assessment of adjunctive ablation strategies, which may have provided additional insights into posterior wall dynamics.

### Clinical relevance of findings

Our findings suggest empirical, isolation of the PW after PVI is not warranted, as the PW may no longer be an important net source of drivers. The ability of GC to characterise the posterior wall as a net sink or source on an individualised basis immediately following PVI positions it as a valuable clinical decision-making tool, potentially guiding real-time recommendations for adjunctive posterior wall isolation in the catheterisation lab for the minority of patients that might benefit. GC or other connectivity mapping may also be able to guide ablation in other areas, including driver ablation, however this requires further evaluation.

## Conclusion

In this study, we describe for the first time the application of GC to global, simultaneous AF mapping data. We found that on average the posterior wall is a net sink following PVI, and therefore PWI will not be beneficial in the majority of patients, providing mechanistic insight into the null randomised control trials for PWI. GC is positioned as a valuable clinical decision tool to select patients that may benefit from PWI to guide personalised ablation strategies in PsAF.

## Data availability statement

The datasets generated or analysed, or both during this study are not publicly available owing to ethical restrictions.

## References

1. Haïssaguerre M, Jaïs P, Shah DC, Takahashi A, Hocini M, Quiniou G, et al. Spontaneous Initiation of Atrial Fibrillation by Ectopic Beats Originating in the Pulmonary Veins. N Engl J Med. 1998;339(10):659–66.

2. Haissaguerre M, Sanders P, Hocini M, Hsu LF, Shah DC, Scavee C, et al. Changes in atrial fibrillation cycle length and inducibility during catheter ablation and their relation to outcome. Circulation. 2004;109(24):3007–13.

3. Drewitz I, Willems S, Salukhe TV, Steven D, Hoffmann BA, Servatius H, et al. Atrial fibrillation cycle length is a sole independent predictor of a substrate for consecutive arrhythmias in patients with persistent atrial fibrillation. Circ Arrhythm Electrophysiol. 2010;3(4):351–60.

4. Badertscher P, Weidlich S, Knecht S, Stauffer N, Krisai P, Voellmin G, et al. Efficacy and safety of pulmonary vein isolation with pulsed field ablation vs. novel cryoballoon ablation system for atrial fibrillation. EP Europace. 2023;25(12).

5. Clarnette JA, Brooks AG, Mahajan R, Elliott AD, Twomey DJ, Pathak RK, et al. Outcomes of persistent and long-standing persistent atrial fibrillation ablation: a systematic review and meta-analysis. EP Europace. 2018;20(FI_3):f366–f76.

6. Sau A, Al-Aidarous S, Howard J, Shalhoub J, Sohaib A, Shun-Shin M, et al. Optimum lesion set and predictors of outcome in persistent atrial fibrillation ablation: a meta-regression analysis. EP Europace. 2019;21(8):1176–84.

7. Jiang X, Liao J, Ling Z, Meyer C, Sommer P, Futyma P, et al. Adjunctive Left Atrial Posterior Wall Isolation in Treating Atrial Fibrillation: Insight From a Large Secondary Analysis. JACC Clin Electrophysiol. 2022;8(5):605–18.

8. William J, Chieng D, Curtin AG, Sugumar H, Ling LH, Segan L, et al. Radiofrequency catheter ablation of persistent atrial fibrillation by pulmonary vein isolation with or without left atrial posterior wall isolation: long-term outcomes of the CAPLA trial. European Heart Journal. 2024.

9. Kaba RA, Momin A, Camm J. Persistent Atrial Fibrillation: The Role of Left Atrial Posterior Wall Isolation and Ablation Strategies. JCM. 2021;10(14):3129.

10. Kistler PM, Chieng D, Sugumar H, Ling L-H, Segan L, Azzopardi S, et al. Effect of Catheter Ablation Using Pulmonary Vein Isolation With vs Without Posterior Left Atrial Wall Isolation on Atrial Arrhythmia Recurrence in Patients With Persistent Atrial Fibrillation: The CAPLA Randomized Clinical Trial. JAMA. 2023;329(2):127–35.

11. Clarke JRD, Piccini JP, Friedman DJ. The role of posterior wall isolation in catheter ablation of persistent atrial fibrillation. J Cardiovasc Electrophysiol. 2021;32(9):2567–76.

12. Tzeis S, Gerstenfeld EP, Kalman J, Saad EB, Sepehri Shamloo A, Andrade JG, et al. 2024 European Heart Rhythm Association/Heart Rhythm Society/Asia Pacific Heart Rhythm Society/Latin American Heart Rhythm Society expert consensus statement on catheter and surgical ablation of atrial fibrillation. EP Europace. 2024;26(4).

13. Calkins H, Hindricks G, Cappato R, Kim YH, Saad EB, Aguinaga L, et al. 2017 HRS/EHRA/ECAS/APHRS/SOLAECE expert consensus statement on catheter and surgical ablation of atrial fibrillation. Heart Rhythm. 2017;14(10):e275–e444.

14. Handa BS, Li X, Aras KK, Qureshi NA, Mann I, Chowdhury RA, et al. Granger Causality–Based Analysis for Classification of Fibrillation Mechanisms and Localization of Rotational Drivers. Circ: Arrhythmia and Electrophysiology. 2020;13(3).

15. Shi R, Parikh P, Chen Z, Angel N, Norman M, Hussain W, et al. Validation of Dipole Density Mapping During Atrial Fibrillation and Sinus Rhythm in Human Left Atrium. JACC Clin Electrophysiol. 2020;6(2):171–81.

16. Grace A, Willems S, Meyer C, Verma A, Heck P, Zhu M, et al. High-resolution noncontact charge-density mapping of endocardial activation. JCI Insight. 2019;4(6).

17. Shi R, Chen Z, Pope MTB, Zaman JAB, Debney M, Marinelli A, et al. Individualized ablation strategy to treat persistent atrial fibrillation: Core-to-boundary approach guided by charge-density mapping. Heart Rhythm. 2021;18(6):862–70.

18. von Elm E, Altman DG, Egger M, Pocock SJ, Gøtzsche PC, Vandenbroucke JP. Strengthening the Reporting of Observational Studies in Epidemiology (STROBE) statement: guidelines for reporting observational studies. Bmj. 2007;335(7624):806–8.

19. Granger CWJ. Investigating Causal Relations by Econometric Models and Cross-spectral Methods. Econometrica. 1969;37(3):424–38.

20. Chen Y, Bressler SL, Ding M. Frequency decomposition of conditional Granger causality and application to multivariate neural field potential data. Journal of neuroscience methods. 2006;150(2):228–37.

21. Claus R, Brandmüller J, Borstel G, Wiesendanger E, Steffan L. Directional dispersion and assignment of optical phonons in LiNbO3. Zeitschrift für Naturforschung A. 1972;27(8-9):1187–92.

22. Deans H. A mathematical model for dispersion in the direction of flow in porous media. Society of Petroleum Engineers Journal. 1963;3(01):49–52.

23. Satyanarayana B, Prasad KS. Discrete mathematics and graph theory: PHI Learning Pvt. Ltd.; 2014.

24. Haissaguerre M, Lim K-T, Jacquemet V, Rotter M, Dang L, Hocini M, et al. Atrial fibrillatory cycle length: computer simulation and potential clinical importance. Europace. 2007;9(Supplement 6):vi64–vi70.

25. Lin Y-J, Tai C-T, Kao T, Chang S-L, Lo L-W, Tuan T-C, et al. Spatiotemporal Organization of the Left Atrial Substrate After Circumferential Pulmonary Vein Isolation of Atrial Fibrillation. Circ: Arrhythmia and Electrophysiology. 2009;2(3):233–41.

26. Lee AM, Aziz A, Didesch J, Clark KL, Schuessler RB, Damiano RJ, Jr. Importance of atrial surface area and refractory period in sustaining atrial fibrillation: testing the critical mass hypothesis. J Thorac Cardiovasc Surg. 2013;146(3):593–8.

